# Optimized tacrolimus dosing strategy in kidney transplant recipients receiving nirmatrelvir-ritonavir for COVID-19

**DOI:** 10.1101/2024.08.22.24312416

**Authors:** Han Yan, Shanbiao Hu, Hedong Zhang, Yangang Zhou, Rao Fu, Ping Xu, Hualin Cai, Xi Li, Gongbin Lan

**Affiliations:** Department of Pharmacy, The Second Xiangya Hospital, Central South University, Changsha Hunan, P.R. China 410011; Institute of Clinical Pharmacy, Central South University, Changsha, China; Department of Kidney Transplantation, The Second Xiangya Hospital, Central South University, Changsha Hunan, P.R. China 410011; Department of Clinical Pharmacology, Xiangya Hospital, Central South University, 87 Xiangya Road, Changsha, Hunan, P.R. China 410008; Institute of Clinical Pharmacology, Central South University; Hunan Key Laboratory of Pharmacogenetics, Changsha Hunan, P.R. China 410008

**Keywords:** COVID-19, tacrolimus, nirmatrelvir-ritonavir, kidney transplant, TDM

## Abstract

Kidney transplantation recipients (KTRs) represent a vulnerable population for COVID-19 infection and severe disease. Nirmatrelvir-ritonavir has demonstrated efficacy in treating COVID-19 among KTRs, and interacts with tacrolimus leading to a precipitous increase in tacrolimus blood levels when co-administered, potentially resulting in toxicity. This study conducted a real-world analysis of KTRs treated with nirmatrelvir-ritonavir for COVID-19 to investigate the relationship between tacrolimus levels and dosing during and within 10 days post-discontinuation of nirmatrelvir-ritonavir. In the experimental group, tacrolimus was initiated at 20-25% of the baseline dose 48 hours after discontinuation of nirmatrelvir-ritonavir, with daily increments of 20-25% until the baseline dose was restored. The patients who did not adhere to the experimental protocol were included in the control group. Findings indicated that withholding tacrolimus 12 hours prior to commencing nirmatrelvir-ritonavir maintained tacrolimus blood levels above 83% of the baseline throughout the nirmatrelvir-ritonavir treatment period. Compared to the control group, the experimental group achieved target trough concentrations of tacrolimus more rapidly and maintained a higher proportion within the therapeutic range (*p*=0.029), and exhibited significantly lower rates of adverse events (*p*<0.001). This investigation provides a safe and effective pharmacological strategy for KTRs infected with COVID-19, enabling the safe co-administration of nirmatrelvir-ritonavir and tacrolimus.

## Introduction

Since the spring of 2020, the world has been experiencing a global COVID-19 pandemic, with the Omicron variant now being the predominant strain spreading widely in communities across various countries(1). Solid organ transplant recipients (SOTRs) are more susceptible to COVID-19 due to long-term immunosuppressant use, which leads to a decline in immune function and a weaker response to vaccines(2). Thus, the virus removal efficiency decreased in SOTRs, and they are at a higher risk of developing severe COVID or death compared to the general population. Multiple studies have demonstrated that SOTR infected with COVID-19 exhibit a significantly elevated mortality rate ranging from 13% to 30%, with a mortality rate of over 50% for those with severe cases (3–8). Therefore, it is crucial for SOTRs to seek prompt treatment when infected with COVID-19 to prevent progression to severe disease (9).

Paxlovid, a medication produced by Pfizer for the treatment of COVID-19, comprises nirmatrelvir and ritonavir. Nirmatrelvir is a novel inhibitor targeting the 3Clpro of SARS-CoV-2 and ritonavir is a potent inhibitor of cytochrome P450 3A, which can reduce the metabolism of nirmatrelvir and elevate its serum levels(10, 11). A phase 2/3 clinical trial indicated that nirmatrelvir-ritonavir reduced the risk of symptomatic COVID-19 progressing to severe disease by 89% compared to the control group (12).

As ritonavir is a potent inhibitor of the CYP3A enzyme system, its interactions with CYP3A-dependent medications may lead to a significant increase in the area under the curve (AUC). Tacrolimus, with a narrow therapeutic window, is commonly used for maintenance anti-rejection therapy after renal transplantation, and any concentration below or exceeding the recommended range could lead to insufficient immunosuppression or toxicity(13, 14). When exposed to ritonavir, the plasma level of tacrolimus will rapidly and significantly increase by approximately 50 times(15). Additionally, after discontinuation of nirmatrelvir-ritonavir, the inhibitory effect on CYP3A enzymes can persist for several days, and resuming the original dose of tacrolimus instantly would lead to a rapid rise in the concentration, potentially causing toxic reactions and increasing the complexity of treatment (16, 17).

Several studies have explored the rational use of nirmatrelvir-ritonavir and tacrolimus in SOTRs. Expert Consensus on the diagnosis and treatment of novel coronavirus infection in SOTRs (2023, China) suggests adjusting the tacrolimus dose by administering a 1/2 daily dose 24 hours after the completion of the nirmatrelvir-ritonavir course, followed by a 3/4 daily dose at 48 hours, and resuming the original dose 72 hours later. However, our patients experienced a significant increase in tacrolimus levels using this scheme, leading to COVID-19 recurrence. Lange *et al.* recommended monitoring tacrolimus levels on the 6th or 7th day of nirmatrelvir-ritonavir initiation and then repeating levels every 2-4 days, holding or restarting with 25% to 75% of the baseline dose based on whether the detected concentration was in the target concentration range (18). However, this frequent monitoring scheme is not suitable for outpatients. Additionally, the activity of the CYP450 enzyme gradually recovers after discontinuation of nirmatrelvir-ritonavir (19), and adjusting the dose only every 2-4 days could result in suboptimal concentrations, potentially increasing the risk of rejection. Therefore, there is an urgent need for further research to determine the optimal timing and dosing regimen for the concurrent use of nirmatrelvir-ritonavir and tacrolimus in renal transplant patients (20–23). Ensuring both the effective treatment of COVID-19 and drug safety is paramount for these patients.

This study conducted a real-world research on renal transplant patients with COVID-19. To explore the relationship between tacrolimus concentration and dosage during nirmatrelvir-ritonavir administration and up to 8 days after drug withdrawal. The findings of this study provide valuable insights into the safe and rational use of tacrolimus in patients with COVID-19 infection after kidney transplantation.

## Methods

### Patient Selection

The data for this study were sourced from the Transplantation Department of the Second Xiangya Hospital, Central South University. We collected clinical data from KTRs who were infected with COVID-19 between January 1, 2023, and July 31, 2023, including follow-up data spanning six months post-infection. All participants had written the informed consent form during data collection. The inclusion criteria were as follows: (i) post-kidney transplantation; (ii) admitted to the Second Xiangya Hospital; (iii) diagnosis of COVID-19 infection with a CT value <30/30 for SARS-CoV-2 nucleic acid; (iv) use tacrolimus for anti-rejection regimen; (v) received nirmatrelvir-ritonavir (Paxlovid, Pfizer) treatment; and (vi) age >18 years and weight >40kg. Patients who were pregnant or breastfeeding, HIV-infected, did not complete the nirmatrelvir-ritonavir treatment course, lacked accessible clinical information, or did not write an informed consent were excluded.

### Therapeutic Regimen

All patients hold tacrolimus 12 hours before starting nirmatrelvir-ritonavir and tacrolimus levels were measured on day 0 (D0). Patients in the experimental group started receiving 20-25% of the baseline dose of tacrolimus 48 hours (D7) after nirmatrelvir-ritonavir withdrawal, 40-50% on D8, 60-75% on D9, 80-100% on D10, and 100% on D11 (day 7 after nirmatrelvir-ritonavir suspension) (17, 19, 23),as shown in Figure 1.

**Fig 1.**
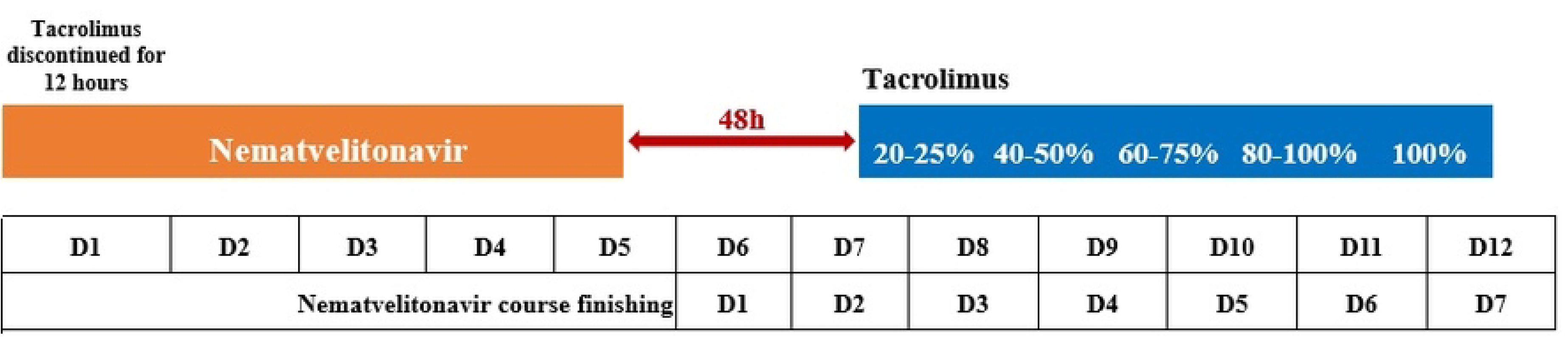
The regimen of commencing tacrolimus after nirmatrelvir-ritonavir withdrawal in the experimental group.

### Samples Grouping

This was a real-world research. Patients who followed the dosing schedule described in Figure 1 were assigned to the experimental group. All patients in the experimental group resumed the original tacrolimus dosage on either the sixth or seventh day after discontinuing of nirmatrelvir-ritonavir. Patients not on this dosing schedule were categorized into the control group, including those who had not resumed the original tacrolimus dosage by the seventh day after stopping nirmatrelvir-ritonavir or had started tacrolimus within 48 hours, as well as those who resumed the original dosage prior to the sixth day. Tacrolimus levels were monitored every 2-3 days during hospitalization, and regular outpatient monitoring of tacrolimus levels and renal function was conducted after discharge.

Information on medication treatment response, including symptom improvement and adverse reactions, was obtained through bedside inquiry or telephone follow-up, while patient outcomes and laboratory test results were retrieved from the medical record system. This study was approved by the ethics committee of the second Xiangya hospital of Central South University (LYEC2024-0016).

### Microbiology

Nasopharyngeal swab testing for SARS-CoV-2 was conducted using real-time fluorescent quantitative RT-PCR technology, targeting the ORF1ab gene and the gene encoding the nucleocapsid protein N for dual-target detection.

### Tacrolimus Level Monitoring

The study included 49 patients with a total of 219 tacrolimus trough concentrations, 41 of which were post-dose data. Tacrolimus level monitoring was performed using Chemiluminescent Microparticle Immunoassay (CMIA) technology on the ARCHITECT i system to quantify FK506 in human whole blood. The test kits were obtained from Abbott Laboratories (United).

### Statistical Analysis

All data in this study were analyzed and plotted using Origin 2020b (Educational Version). The cumulative survival curve was plotted using the Kaplan-Meier method, and the log-rank test was used to assess significant differences in cumulative risk. Independent sample t-tests and chi-square tests were used to compare significant differences between the numerical and categorical variables, respectively. All tests were two-tailed, with *p*<0.05 considered statistically significant.

## Results

### Baseline Characteristics

A total of 49 hospitalized kidney transplantation patients who met the criteria for COVID-19 infection admitted to the Second Xiangya Hospital were enrolled. The baseline characteristics of the cohort are presented in Table 1. In brief, the median age was 47 years (range 18-65), and 30 patients (61.2%) were male. The median time from transplantation to COVID-19 diagnosis was 44 months (range 0-240). Among them, there were 8 patients (16.3%) with diabetes, 21 (42.9%) with hypertension, 4 (8.2%) with osteoporosis, and 21 (42.9%) with anemia. Immunosuppressive treatment mainly included a combination of tacrolimus (100%), mycophenolate mofetil (93.9%), and steroids (100%). 19 patients (38.8%) received medium-dose steroid anti-inflammatory treatment. Only 9 patients (18.4%) and 7 patients (14.3%) were vaccinated with three doses and two doses, respectively. None of the patients had a history of Sars-CoV-2 infection, and the median time from symptom onset to hospital admission was 4 days (range 0-30 days). The symptoms after infection included fever (73.4%), cough (67.3%), fatigue (40.8%), muscle and throat pain (38.8%), and chest tightness and shortness of breath (28.6%). All patients completed 5 days of nirmatrelvir-ritonavir therapy, with dosage adjustments as follows: GFR >30ml/min/1.73m^2 given 300/100mg q12h, GFR between 10-30ml/min/1.73m^2 given 150/100mg q12h, GFR <10ml/min/1.73m^2 given 150/100mg qd.

**Table 1.**
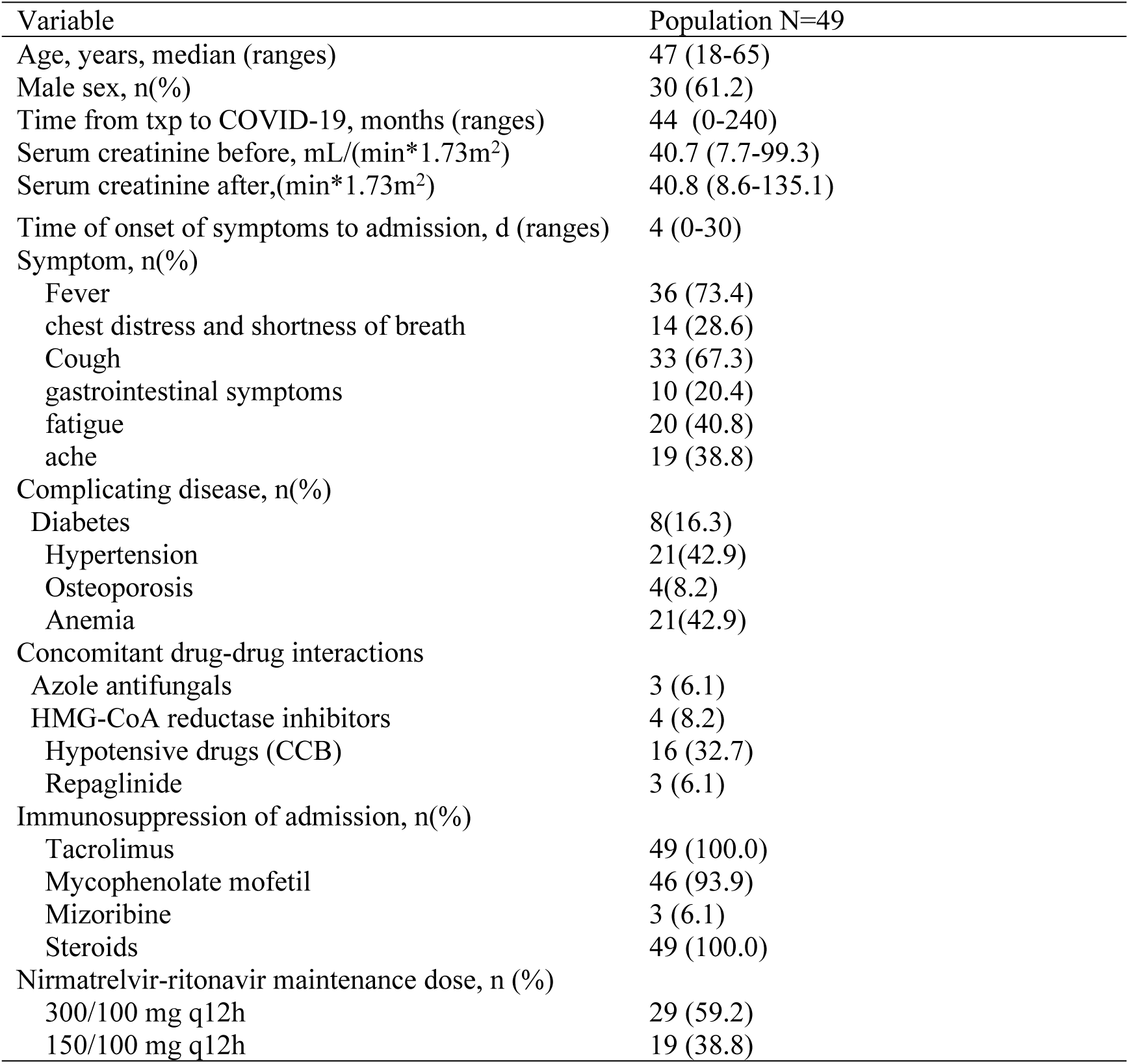
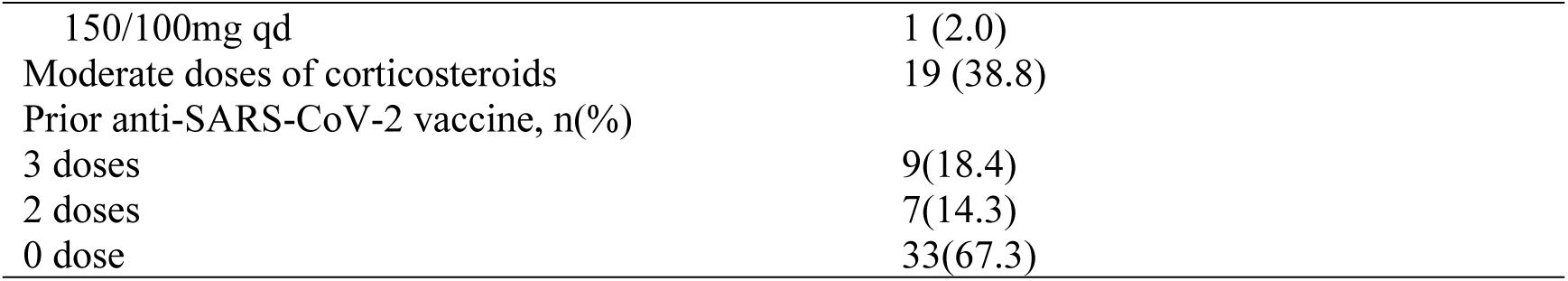
Baseline characteristics of the cohort.

| Variable | Population N=49 |
| --- | --- |
| Age, years, median (ranges) | 47 (18-65) |
| Male sex, n(%) | 30 (61.2) |
| Time from txp to COVID-19, months (ranges) | 44 (0-240) |
| Serum creatinine before, mL/(min*1.73m<sup>2</sup>) | 40.7 (7.7-99.3) |
| Serum creatinine after,(min*1.73m<sup>2</sup>) | 40.8 (8.6-135.1) |
| Time of onset of symptoms to admission, d (ranges) | 4 (0-30) |
| Symptom, n(%) |  |
| Fever | 36 (73.4) |
| chest distress and shortness of breath | 14 (28.6) |
| Cough | 33 (67.3) |
| gastrointestinal symptoms | 10 (20.4) |
| fatigue | 20 (40.8) |
| ache | 19 (38.8) |
| Complicating disease, n(%) |  |
| Diabetes | 8(16.3) |
| Hypertension | 21(42.9) |
| Osteoporosis | 4(8.2) |
| Anemia | 21(42.9) |
| Concomitant drug-drug interactions |  |
| Azole antifungals | 3 (6.1) |
| HMG-CoA reductase inhibitors | 4 (8.2) |
| Hypotensive drugs (CCB) | 16 (32.7) |
| Repaglinide | 3 (6.1) |
| Immunosuppression of admission, n(%) |  |
| Tacrolimus | 49 (100.0) |
| Mycophenolate mofetil | 46 (93.9) |
| Mizoribine | 3 (6.1) |
| Steroids | 49 (100.0) |
| Nirmatrelvir-ritonavir maintenance dose, n (%) |  |
| 300/100 mg q12h | 29 (59.2) |
| 150/100 mg q12h | 19 (38.8) |
| 150/100mg qd | 1 (2.0) |
| Moderate doses of corticosteroids | 19 (38.8) |
| Prior anti-SARS-CoV-2 vaccine, n(%) |  |
| 3 doses | 9(18.4) |
| 2 doses | 7(14.3) |
| 0 dose | 33(67.3) |

### Clinical Efficacy and Adverse Drug Reaction of Nirmatrelvir-ritonavir

All 49 patients with COVID-19 infection recovered after treatment with nirmatrelvir-ritonavir. Sixteen patients tested negative for COVID-19 nucleic acid after completing the treatment course (5 days), and the remaining patients turned antigen negative or symptoms disappeared within two weeks after discharge. Additionally, two patients with COVID-19 rebounded after one course of nirmatrelvir-ritonavir treatment, and two patients received two courses of nirmatrelvir-ritonavir treatment due to unresolved symptoms. After treatment, the SARS-CoV-2 Ct value increased significantly (*p*<0.001), indicating a reduction in the viral load (Figure 2). During treatment, 40 patients (81.6%) experienced symptoms such as bitter taste and altered taste sensation, 13 patients (26.5%) experienced gastrointestinal discomfort, 2 patients (4.1%) experienced dizziness, and 1 patient (2.0%) had itchy hands. These adverse reactions were alleviated after discontinuation of medication.

**Fig 2.**
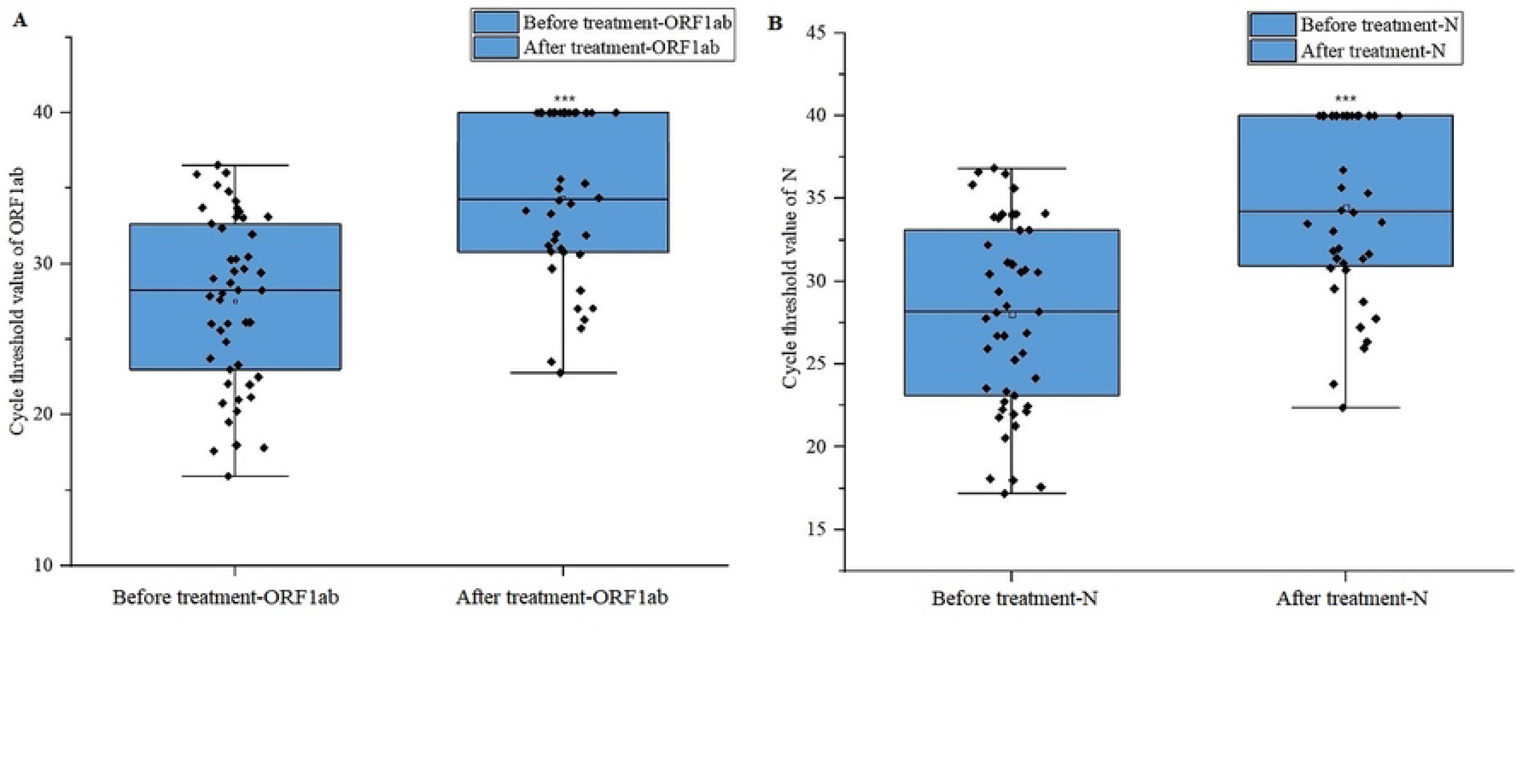
Changes in the SARS-CoV-2 Ct value in patients before and after nirmatrelvir-ritonavir administration. A: ORF1ab gene. B: The gene encoding the nucleocapsid protein N. *** represent p<0.001.

### Tacrolimus Concentration Changes During Nirmatrelvir-ritonavir Use

All patients discontinued tacrolimus 12 hours before starting nirmatrelvir-ritonavir treatment, and the trough concentration of tacrolimus measured before medication was used as the baseline concentration. Tacrolimus concentrations were monitored every 2-3 days, and 219 blood drug concentration measurements were collected. After removing 41 post-dose data, there were 178 TDM data during the tacrolimus discontinuation period in this study. The results showed that despite the withdrawal of tacrolimus, its concentration could be maintained at a relatively high level during the administration of nirmatrelvir-ritonavir, with D1-D5 trough concentrations of 98%, 107%, 96%, 83%, and 89% of the baseline concentration, respectively. On D6 and D7, tacrolimus concentrations could still be maintained at 69% and 60%, respectively, while on D8 and thereafter, its blood drug concentration dropped significantly to lower levels of 47%, 30%, 26%, None, 16%, and 10%, respectively (Figure 3). Therefore, within 48 hours of ritonavir discontinuation, tacrolimus concentration could still be maintained at a relatively high level.

**Fig 3.**
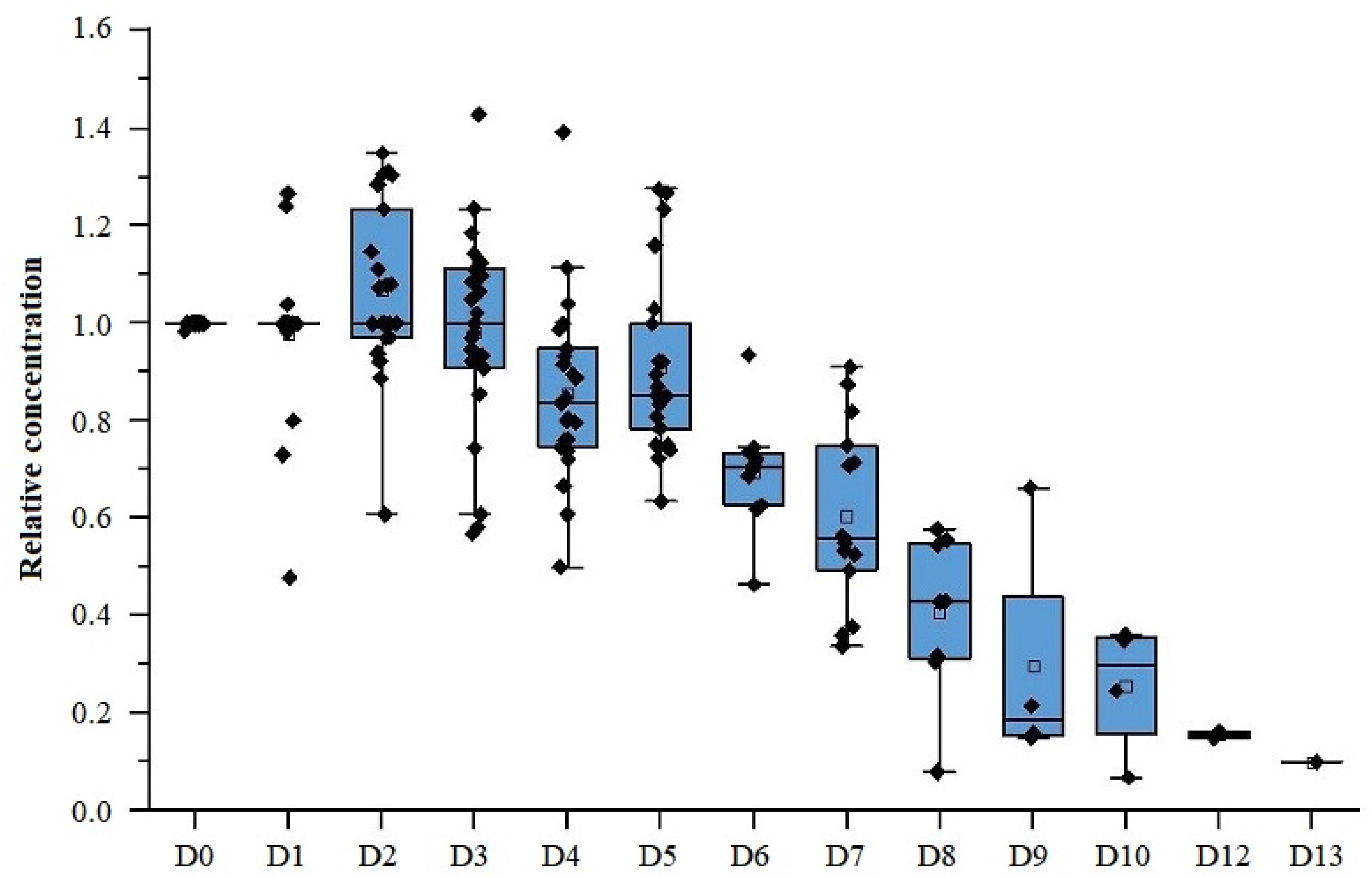
Changes of residual blood concentration of tacrolimus during and after nirmatrelvir-ritonavir use. The TDM data presented in this figure was exclusively collected during the periods when the subjects were not administering tacrolimus due to other reasons such as secondary severe infection or unrelieved symptoms. Note: There is no TDM data on day 11.

### Exploration of Tacrolimus Dosing Regimen After Nirmatrelvir-ritonavir Discontinuation

This part of the study included 34 patients who resumed tacrolimus treatment after the end of nirmatrelvir-ritonavir treatment and underwent TDM monitoring within 10 days at the Second Xiangya Hospital. After excluding 9 samples who used Wuzhi Capsules (CYP3A enzyme inhibitors, which could increase the oral bioavailability of tacrolimus and increase its blood concentration) together(24), 11 patients were included in the experimental group and 14 in the control group, with a total of 69 FK506 concentration tests conducted. Concentration compliance was defined as fluctuations within 20% of baseline concentration. Due to limited data, concentrations measured every two days were grouped together for analysis. The ratio in the target concentration range of tacrolimus in the experimental group was significantly higher than that in the control group (*p*=0.029) (Figure 4A), and the cumulative compliance ratio analysis showed that the experimental group patients reached the target range faster and with a higher proportion (72.7% vs 21.4%, *p*=0.001) (Figure 5). Additionally, concentrations in the experimental group that fell outside the target range were still close to the target concentration, whereas concentrations in the control group deviated further from the target range (Figure 4B).

**Fig 4.**
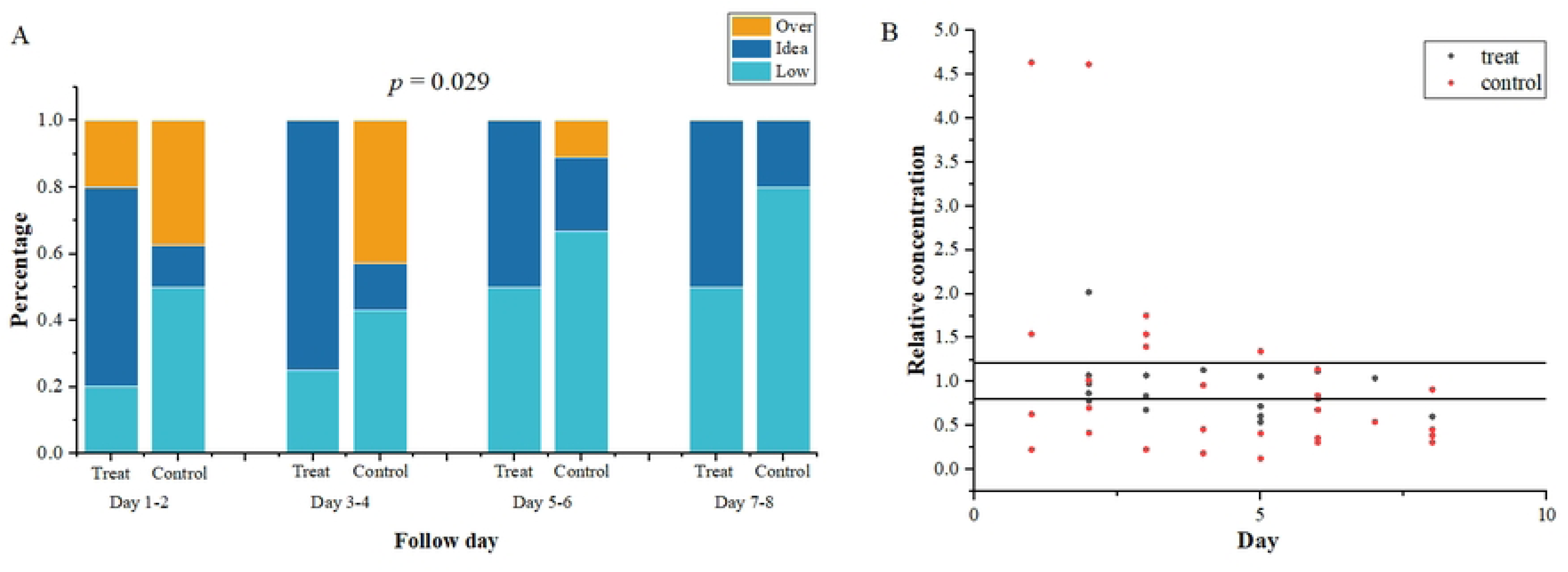
The ratio in the target concentration range of tacrolimus between experimental group and control. A: Ratio of reaching the target concentration range in every two days. B: Scatter plot of relative concentration of tacrolimus.

**Fig 5.**
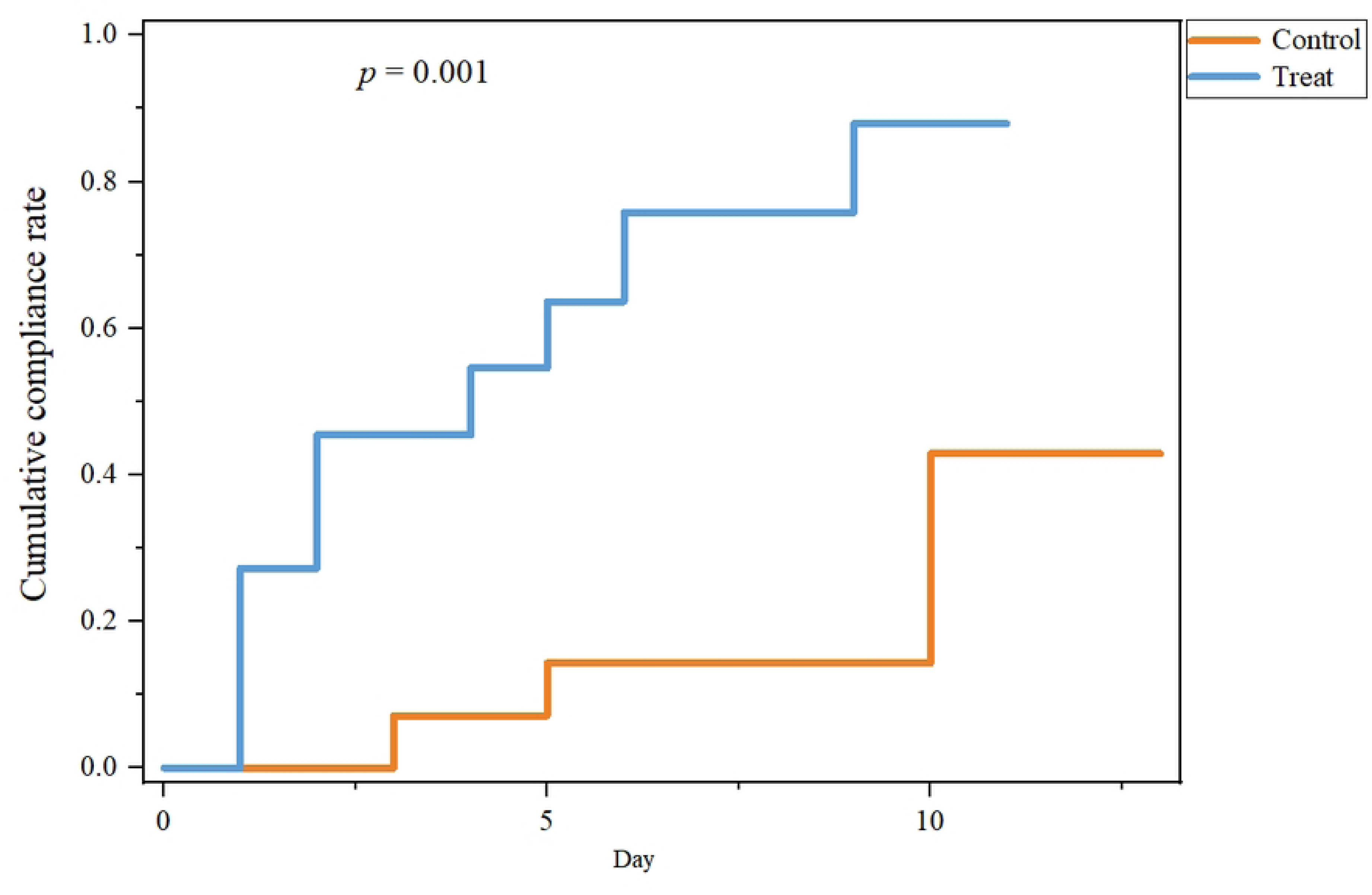
The cumulative compliance ratio analysis of tacrolimus between experimental group and control.

In addition, two patients in the control group began to take 80%, 45% of the basic dose of tacrolimus within 12 hours after the withdrawal of nirmatrelvir-ritonavir, followed by 40% and 100% the next day. Then their concentration significantly increased to more than 4.64 and 4.62 times of the baseline concentration, with 5.5 ng/ml rising to 26 ng/ml, and 6.5 ng/ml rising to more than 30ng/ml. Another patient took 22% of the baseline dose of tacrolimus for two consecutive days after nirmatrelvir-ritonavir discontinuation for 24 hours, and the concentration increased to 2.02 times of the baseline concentration. However, for patients who did not resume tacrolimus 48 hours after nirmatrelvir-ritonavir withdrawal, their concentration was significantly lower than the baseline concentration (Figure 3). Patients who received insufficient doses compared to the experimental group also had significantly lower concentrations than the baseline concentration.

Furthermore, we encountered a special case in the clinical practice that was not included in this study. One patient did not follow medical advice and self-administered tacrolimus with 1mg q12h on the third and fourth days of nirmatrelvir-ritonavir administration, causing the tacrolimus concentration to rise from 2.0ng/ml to >30ng/ml, and the serum creatinine from 98μmol/L to 123μmol/L, with noticeable tremors occurring. Tacrolimus was immediately discontinued, and at this point, nirmatrelvir-ritonavir was completed. Two days later, the tacrolimus concentration was still over 30ng/ml. Over the next two days, CYP3A4 enzyme inducers rifampin 450mg and 300mg were administered for the following two days, respectively. Subsequently, the patient’s tacrolimus concentration dropped to 8.3ng/ml. One week after discharge, the patient was followed-up for negative detection of the COVID-19 antigen, and renal function recovered to pre-admission levels.

### Events Occurring Within 6 Months After COVID-19 Recovery

In this part of the study, two patients were treated with nirmatrelvir-ritonavir twice each for COVID-19 positive. Therefore, the number of patients in the control and treat group was 13 and 10, respectively. Among the 13 patients in the control group, 3 were hospitalized due to creatinine elevation within 6 months, 4 had COVID-19 rebound or delayed conversion to negative, 2 experienced opportunistic infections, one with Pneumocystis jiroveci pneumonia (PJP) and the other with Cytomegalovirus (CMV), and one patient died from severe pneumonia triggered by herpes virus infection. The event rate within 6 months in the control group was 69%, while none of the patients in the experimental group experienced the aforementioned events. The incidence of events within 6 months after nirmatrelvir-ritonavir treatment for COVID-19 was significantly lower in the experimental group than in the control group (69% vs 0%, *p*<0.001) (Table 2).

**Table 2.** Events occurred within 6 months after treatment of COVID-19 infection with nirmatrelvir-ritonavir.

| Events | Control (N=13) | Treat (N=10) | P value |
| --- | --- | --- | --- |
| Admitted with Creatinine elevation, n(%) | 3(23.1) | 0(0) |  |
| COVID-19 rebounded or negative conversion time prolonged, n(%) | 4(30.8) | 0(0) |  |
| Opportunistic infections, n(%) | 2(15.4) | 0(0) |  |
| All-cause mortality, n(%) | 1(7.7) | 0(0) |  |
| Total, n(%) | 9(69.2)# | 0(0) | *p*<0.001 |

#: One patient has both COVID-19 rebounded and *Pneumocystis jiroveci* pneumonia infection.
Note: Among the samples in the study, two patients were treated with nirmatrelvir-ritonavir twice for COVID-19 positive. Thus the number of patients in control group and treat group were 13 and 10, respectively.

## Discussion

This Real-world study focused on patients with COVID-19 after kidney transplantation. Notably, the administration of nirmatrelvir-ritonavir exhibited remarkable efficacy in this patient population, achieving a 100% clinical cure with minimal adverse reactions. Although tacrolimus was withheld for 12 hours prior to the commencement of nirmatrelvir-ritonavir therapy, tacrolimus concentration remained above 83% of the baseline level during the ritonavir administration. At 48 hours after the cessation of ritonavir, commencing tacrolimus at a low dose of 20-25% of the baseline, with daily increments of 20-25% of the baseline dose, allowed for faster and more consistent maintenance of the concentration of tacrolimus within the desired range. Remarkably, compared to the control group, patients in the experimental group exhibited significantly lower rates of events such as COVID-19 rebounded or delayed conversion to negative, hospitalization due to creatinine elevation, opportunistic infections, and all-cause mortality in the subsequent six-month follow-up period. The medication strategy proposed in this study necessitated patients to detect blood concentration once only 24 hours after re-initiating tacrolimus and determine the precise timing for restoring the original dosage, either on D6 or D7, based on whether it is within the target concentration range, making medication more concise and safe for patients.

Renal transplant patients are at high risk of COVID-19 infection and developing severe disease. Currently, the most effective drugs for Omicron infection mainly include nirmatrelvir-ritonavir, *etc.* Nirmatrelvir is a 3CLpro inhibitor that alleviates disease progression by blocking the replication of coronavirus. It is a substrate of CYP3A, and ritonavir is a potent, irreversible inhibitor of CYP3A4 and also an inhibitor of the P-glycoprotein transporter. Combined use can increase the blood concentration of nirmatrelvir, thereby enhancing its efficacy. Tacrolimus is a substrate of CYP3A4/5 and is transported by P-glycoprotein. When used in combination with ritonavir, it can lead to a significant increase in concentration, resulting in elevated serum creatinine levels, aggravated infections, and even COVID-19 re-positivity, posing a significant threat to patient safety(25). In our research, we also observed similar clinical phenomenon, as mentioned in the Results section. Some patients experienced a distinct increase in creatinine levels due to patients not following medical advice and self-administering tacrolimus, which gradually returned to pre-admission levels after discontinuing the medication.

Furthermore, although the half-life of ritonavir is approximately 5.6 hours, its inhibitory effects on CYP3A4 and 3A5 persists after discontinuation. Studies have shown that CYP3A activity reaches nearly 75% of metabolic activity after 48 hours of ritonavir withheld, and fully recovered at 2-5 days (18, 19, 26). Our results showed that 48 hours after discontinuation of ritonavir, the tacrolimus concentration, which had been discontinued for seven days, could still reach 60.5% of the baseline concentration. This suggests that adding tacrolimus before this point could lead to a sharp increase in its concentration.

In our study, patients in the experimental group discontinued tacrolimus 12 hours prior to initiating nirmatrelvir-ritonavir therapy, and 48 hours after nirmatrelvir-ritonavir withdrawal, they began administering tacrolimus at 20-25% of the original dose, gradually increasing daily. By the sixth to seventh days of nirmatrelvir-ritonavir discontinuation, all patients had resumed their original tacrolimus dosage. Although the blood concentration of FK506 gradually decreased following tacrolimus suspended, reaching approximately 60% of the baseline concentration on the seventh day. Due to the temporary discontinuation of the medication, there was no peak concentration, resulting in an even lower AUC for tacrolimus. However, multiple studies have shown that after SARS-CoV-2 invades the human body, it impairs the host immune system in various ways, leading to a reduction or even exhaustion of T cells (27–34). Consequently, patients experience a weakened immune status during COVID-19 infection, reducing the risk of acute kidney transplant rejection. Therefore, there is no urgent need to resume tacrolimus therapy prematurely to maintain the pre-infection AUC levels. Furthermore, excessive efforts to achieve baseline tacrolimus concentration levels during this period may hinder virus clearance, delay recovery, and even increase the risk of rebounded (35). Consistent with this, our follow-up data revealed that even when tacrolimus was discontinued during nirmatrelvir-ritonavir therapy and gradually added at low doses, none of the patients experienced acute rejection during the 6-month follow-up period. Conversely, patients who prematurely resumed tacrolimus exhibited delayed COVID-19 negativity or recurrence and an increased risk of opportunistic infections. Nonetheless, for patients with high immunological risk factors, such as those with recent transplants or a history of rejection, clinicians may strongly consider allowing patients to achieve low tacrolimus concentrations on the third day of nirmatrelvir-ritonavir treatment to avoid suboptimal treatment levels. Alternatively, they may opt to restart low-dose tacrolimus immediately after completing nirmatrelvir-ritonavir therapy or adopt a close monitoring and level-based dosing approach (36).

This study had some limitations. First, it focuses on patients who have been in a stable condition for one month or longer following kidney transplantation. Consequently, the research has not delved into how to adjust tacrolimus dosing for patients with high immunological risk factors, such as those who have undergone recent transplantation or have a history of rejection. Second, we do not examine the safe administration of tacrolimus in conjunction with CYP3A enzyme inhibitors, including Wuzhi Capsules. Third, this is a single-center small-sample real-world study, and the findings require further validation through multi-center and expanded samples.

## Conclusion

Our findings present a refined and standardized dosing regimen that mitigates the interaction between nirmatrelvir-ritonavir and tacrolimus. The judicious use of nirmatrelvir-ritonavir can significantly aid renal transplant recipients in treating COVID-19 infection more effectively, thereby shortening the disease duration and minimizing the occurrence of severe cases. The gradual introduction of tacrolimus with daily increments of 20-25% of the baseline dose, commencing 48 hours after the cessation of nirmatrelvir-ritonavir, can reduce the risk of COVID-19 recurrence and opportunistic infections, while maintaining a low risk of allograft rejection. This approach ensures the medication safety for renal transplant patients after COVID-19 infection.

## Acknowledgments

This research was supported by the Chinese National Science Foundation (No.81803583) and Hunan Provincial Natural Science Foundation of China (2019JJ50854, 2021JJ31074 and 2023JJ30755).

## Authors’ contributions

Han Yan and Gongbin Lan designed the study. Han Yan wrote the manuscript. Gongbin Lan, Shanbiao Hu, Hedong Zhang, Rao Fu and Hualin Cai participated in sample collection. Yangang Zhou and Ping Xu made some suggestions for this research. Xi Li and Han Yan analyzed the data and prepared the figures. All authors have reviewed the manuscript.

## Disclosure

### Ethics approval and consent to participate

This study was approved by the Ethics Committee of the Institute of Clinical Pharmacology, Central South University. None of the donor organs for the patients in this study were procured from executed prisoners, and the organs were procured after informed consent or authorization.

## Data availability statement

The data that support the findings of this study is available from the corresponding author upon a reasonable request.

## Abbreviation

KTR: Kidney transplantation recipient
COVID-19: Coronavirus disease 2019
TDM: Therapeutic drug monitoring
SOTR: Solid organ transplant recipient
CYP450: Cytochrome P450
SARS-CoV-2: Severe acute respiratory syndrome coronavirus 2
AUC: Area under the curve
CMIA: Chemiluminescent Microparticle Immunoassay
GFR: glomerular filtration rate
CMV: Cytomegalovirus
PJP: Pneumocystis jiroveci pneumonia
3CLpro: 3C-like protease

